# Exploring real-world concordance of tissue mutation burden (TMB) from blood and tissue in patients with solid tumors

**DOI:** 10.1101/2021.05.05.21256698

**Authors:** Jewel Park, Inae Park, Jin Young Hwang, William Han Bae, Myungwoo Nam, Grace Lee, Leeseul Kim, Yoonhee Choi, Hojung Jung, Min Jeong Kim, Seung Pyo Daniel Hong, Hiwon Lee, Younggyeong Park, Emma Yu, Alice Lee, Young Kwang Chae

**Author notes:** Corresponding author: Young Kwang Chae, M.D., M.P.H., M.B.A, 645 N. Michigan Avenue, Suite 1006, Chicago, IL 60611, |. Disclaimers: Dr. Young Kwang Chae receives research grant from Abbvie, BMS, Biodesix, Lexent Bio, and Freenome. He has received honoraria from or is in advisory boards for Roche/Genentech, BMS, AstraZeneca, Merck, Foundation Medicine, Counsyl, Neogenomics, Guardant Health, Boehringher Ingelheim, Biodesix, Immuneoncia, Lilly Oncology, Merck, Takeda, Pfizer, Tempus, Lunit, and Jazz Pharmaceuticals.

## Abstract

**Purpose:** Tumor mutational burden (TMB) is an approved biomarker for immunotherapy in metastatic cancer patients. While initially measured from tissue (tTMB), TMB derived from circulating tumor DNA (ctDNA) - also known as blood TMB (bTMB) - is increasingly being used in the clinic. Currently, real-world concordance between tTMB and bTMB is not well understood.

**Patients and methods:** From October 2020 to March 2021, cancer patients who had both tTMB and bTMB results were selected. Patients were classified according to clinical variables and tumor burden, and correlation analyses or tests of independence were performed to explore any associations.

**Results:** From a total of 38 patients included in our study, 20 patients (52.6%) had non-small cell lung carcinoma and 18 (47.4%) had other cancers. Median bTMB of 9.6 mut/MB was higher than median tTMB of 4.0 mut/Mb, and the distributions of bTMB and tTMB differed significantly (n=38, p < 0.001). bTMB was positively correlated with tTMB in the total study population (Spearman ρ=0.57, p < 0.001) and a tTMB of 10 mut/Mb correlated with a bTMB of 21.1 mut/Mb. Dividing patients by cancer type or site of tumor biopsy resulted in significantly differing strength and degree of correlation, but dividing patients by concordant and discordant bTMB:tTMB ratio did not reveal any significantly different distributions of clinical variables or tumor burden.

**Conclusion:** bTMB was positively correlated with tTMB, and median bTMB was higher than median tTMB. Cancer type and site of tissue biopsy may influence concordance between tTMB and bTMB. Future studies with more patients may help define the optimal bTMB threshold for receiving immunotherapy, which may be different from the tTMB threshold.

## Introduction

Tumor mutation burden (TMB) is a predictor of response to immunotherapy in diverse cancers.^1^ In June 2020, pembrolizumab was approved by the United States Food and Drug Administration for the treatment of unresectable or metastatic solid tumors with TMB ≥10 mutations/megabase (mut/Mb) based on the data from the KEYNOTE-158 trial, which used TMB derived from tissue, i.e. tissue TMB (tTMB).^2^ Recent studies have tried to characterize TMB from blood instead of tissue, since collecting blood samples is less invasive, more amenable to monitoring tumor evolution, and more likely to better reflect tumor heterogeneity.^3^

The extent to which bTMB is concordant with tTMB is unclear. Numerous trials have suggested that bTMB can also be used as a predictor of favorable response to immunotherapy, and that bTMB levels correlate with tTMB levels at a ratio of approximately 1.6:1.^4–6^ However, contrasting results have been reported as well, where bTMB only weakly correlated with tTMB and was even associated with shorter overall survival.^7, 8^ This may be in part due to the lack of harmonization among currently available targeted gene platforms for TMB calculation, since each platform covers different numbers of genes and coding regions.^9^

Patient characteristics and clinical variables may also contribute to the discordance between bTMB and tTMB. Previous reports have suggested that variables such as smoking history, tumor burden, and time interval between tissue and blood sampling can be confounding factors.^10–12^ Moreover, tTMB measured from metastatic tumors are higher than that from primary tumors, but this is often overlooked when comparing bTMB and tTMB.^13^

Tumor mutational burden and its measurement is increasingly being incorporated into routine practice to guide therapy in cancer patients.^9^ However, there is a concern that immunotherapy can be used on patients inappropriately, especially since the concordance between bTMB and tTMB in real-world settings is not yet clear. Thus, we attempted to explore this relationship using real-world data from two commercially available platforms that measure tTMB and bTMB.

## Methods

### Patient selection and study design

. The study was approved by the Institutional Review Board (IRB) of Northwestern University Feinberg School of Medicine. Since retrospective review of molecular analyses was performed, informed consent was waived. Studies were performed in concordance with the Health Insurance Portability and Accountability Act and the Declaration of Helsinki. Patients treated at the Robert H. Lurie Comprehensive Cancer Center of Northwestern University from October 2020 to March 2021 were retrospectively identified to have commercial NGS testing for both tissue and blood, i.e. by Tempus xT (Tempus; Chicago, IL) and Guardant360 (Guardant Health; Redwood City, CA), respectively. From this cohort, only patients who were treatment-naïve or treatment-refractory at the time of blood sample collection were included for final analysis, as bTMB levels of patients who were responding to treatment may have been significantly altered by therapy. To explore the significance of concordant and discordant bTMB:tTMB ratio, patients were divided into tertiles based on their bTMB:tTMB ratio.The first and third tertiles were defined as “low” and “high”, respectively, while the second tertile was defined as “mid”. “Low” and “high” subgroups were considered to be discordant while “mid” was considered to be concordant.

### Next-generation sequencing and TMB calculation

All patients in the study had NGS testing performed by both Tempus xT and Guardant360. Tempus xT assay consists of 648 genes with single nucleotide variants (SNV), indels, and translocations measured by hybrid capture NGS. In Guardant360, the TMB score is calculated from somatic SNVs and indels in exons of ~ 500 genes detected in cell-free DNA, followed by adjusting for tumor shedding levels and the size of the panel.

### Measuring tumor burden

Tumor burden was assessed using imaging and NGS of blood samples. The size of tumors from CT and PET-CT images were assessed by independent radiologists and calculated to give a final score according to the Response Evaluation Criteria In Solid Tumors version 1.1 (RECIST v1.1). The maximum allele frequency (MAF), which is a measure of the highest frequency clone, is provided in the Guardant360 report and was used as a molecular marker of tumor burden.

### Statistical analysis

The TMB levels of our study population had a non-parametric distribution, so Spearman’s test was used to assess linear correlation and Wilcoxon’s or Kruskal-Wallis test was used to compare median values and distributions of patient subgroups. Chi-square test was used to compare frequency distributions of patients divided into tertiles by bTMB:tTMB ratio. All analyses were performed using R version 4.0.4.

## Results

### Patient characteristics

Table 1 shows the baseline characteristics of the 38 patients in our study. The median age was 67 years. Lung adenocarcinoma was the most common cancer type with 13 patients, followed by lung squamous cell carcinoma with seven patients; other types of cancer included small cell lung, esophageal, gastric, appendiceal, pancreatic, breast, ovarian, cervical, endometrial, uterine, thymic, thyroid, and maxillary cancer. Tissue biopsy was taken from primary lesions in 18 patients, from lymph nodes in nine patients, and other metastatic lesions in ten patients.

**Table 1.** Baseline patient characteristics of 38 patients.

| **Characteristic** | **N** | **Percentage** |
| --- | --- | --- |
| Age, median (range) | 67 (31-90) |  |
| Gender |  |  |
| Female | 20 | 52.6 |
| Male | 18 | 47.4 |
| Cancer type |  |  |
| Lung adenocarcinoma | 13 | 34.2 |
| Lung squamous cell carcinoma | 7 | 18.4 |
| Other cancers | 18 | 47.4 |
| Stage |  |  |
| I/II/III | 4 | 10.5 |
| IV | 34 | 89.5 |
| Site of tissue biopsy |  |  |
| Primary tumor | 19 | 50.0 |
| Metastatic tumor | 10 | 26.3 |
| Lymph node | 9 | 23.7 |
| Sampling interval |  |  |
| <30 days | 17 | 44.7 |
| ≥30 days | 21 | 55.3 |
| Smoking history |  |  |
| Never smoker | 9 | 23.7 |
| Former/current smoker | 29 | 76.3 |

### Correlation between bTMB and tTMB

Median values and interquartile range of tissue and blood TMB are shown in Table 2. Median bTMB of 9.6 mut/MB was higher than median tTMB of 4.0 mut/Mb, and the distributions of bTMB and tTMB differed significantly (Wilcoxon signed-rank V=14.5, n=39, p<0.001). bTMB was moderately correlated with tTMB (Spearman ρ=0.56, p < 0.001), and a bTMB of 21.1 mut/Mb correlated with tTMB of 10 mut/Mb (Figure 1). Two patients had tTMB ≥ 10 mut/Mb while 16 patients had bTMB ≥ 10 mut/Mb. When patients were divided according to cancer type, correlation between bTMB and tTMB was not statistically significant for both lung adenocarcinoma and lung squamous cell carcinoma, and regression lines for both lung cancer subtypes displayed marked differences in slope. Furthermore, dividing patients by site of tissue biopsy revealed that the degree of correlation was pronounced for tissue samples from metastatic sites (ρ = 0.88, p < 0.001).

**Table 2.** Summary of tissue and blood tumor mutational burden levels.

|  | Tissue TMB |  |  |  | Blood TMB |  |  |  |
| --- | --- | --- | --- | --- | --- | --- | --- | --- |
|  | 1st Qu. | Median | Mean | 3rd Qu. | 1st Qu. | Median | Mean | 3rd Qu. |
| All cancers | 2.6 | 4.0 | 4.9 | 6.2 | 7.8 | 9.6 | 12.5 | 12.3 |
| NSCLC | 2.6 | 4.2 | 5.2 | 6.4 | 7.6 | 9.5 | 13.7 | 15.6 |
| Other cancers | 2.8 | 3.2 | 4.7 | 5.3 | 8.2 | 9.6 | 11.0 | 10.5 |

TMB, tumor mutational burden; NSCLC, non-small cell lung cancer.

**Figure 1.**
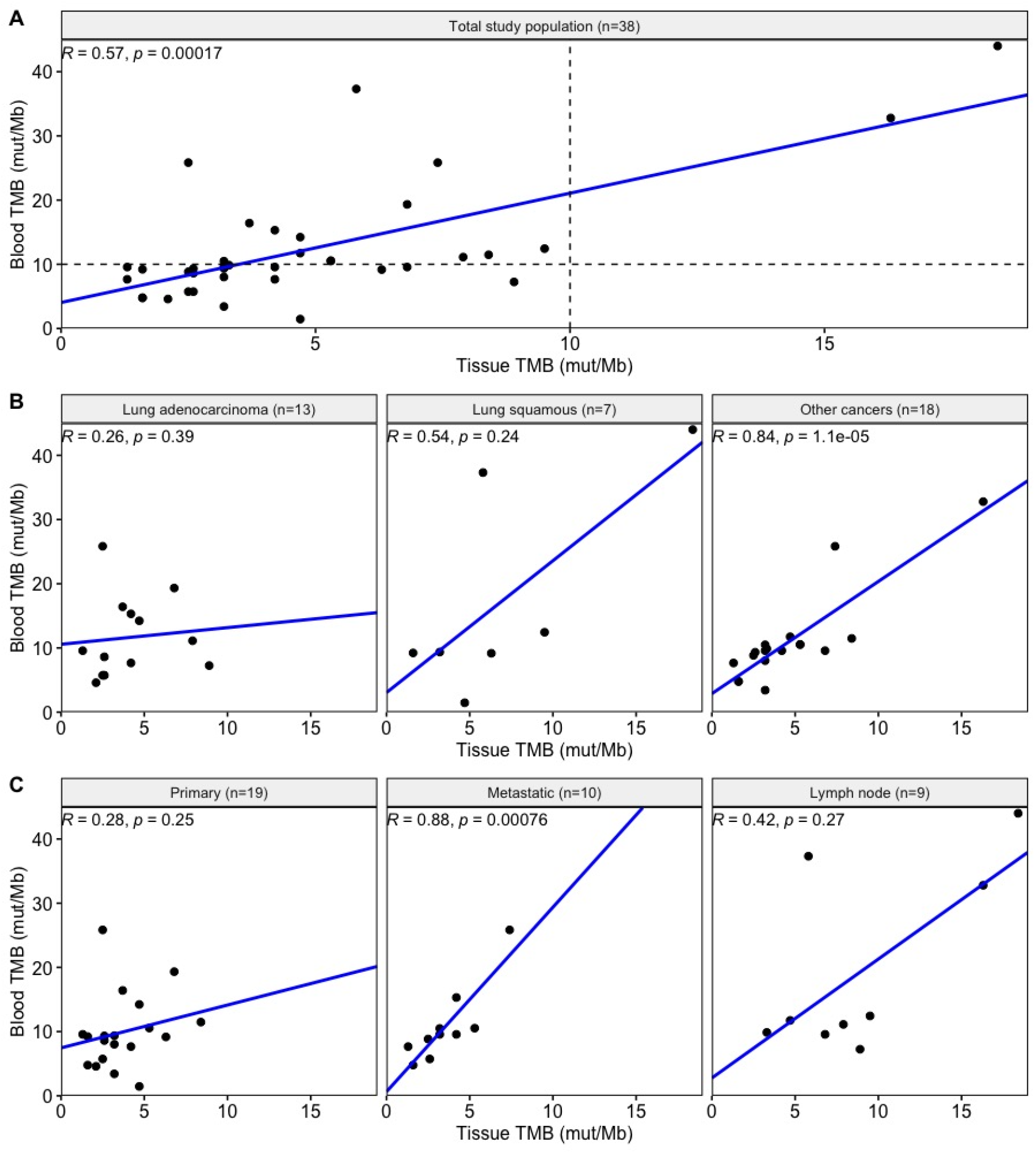
Correlation between tumor mutational burden from blood and tissue Spearman correlations were examined in A) the total study population, B) patients divided by cancer type, and C) patients divided by site of tissue biopsy. Dotted line in Figure 1A represent the >10 mut/Mb cutoff for TMB. TMB, tumor mutational burden.

### Distribution of patients according to bTMB:tTMB ratio

Patients were divided by their bTMB:tTMB ratio into tertiles of “low” (0-2.2), “mid” (2.2-3.0), and “high” (3.0-10.3). No significant differences in frequency distribution were observed for any of the patient characteristics, such as gender, cancer type, site of tissue biopsy, sample interval, and smoking status, or measures of tumor burden (Table 3).

**Table 3.** Distribution of patient characteristics and tumor burden according to bTMB:tTMB ratio divided into tertiles.

|  | **Low (0.0-2.2)** | **Mid (2.2-3.0)** | **High(3.0-10.3)** | **P-value** |
| --- | --- | --- | --- | --- |
| *Patient characteristics* |  |  |  |  |
| Gender |  |  |  | 0.666 |
| Female | 6 | 6 | 8 |  |
| Male | 7 | 6 | 5 |  |
| Cancer type |  |  |  | 0.800 |
| Lung adenocarcinoma | 4 | 3 | 6 |  |
| Lung SCC | 3 | 2 | 2 |  |
| Other cancers | 6 | 7 | 5 |  |
| Site of tissue biopsy |  |  |  | 0.238 |
| Primary tumor | 7 | 5 | 7 |  |
| Metastatic tumor | 1 | 4 | 5 |  |
| Lymph node | 5 | 3 | 1 |  |
| Sampling interval |  |  |  | 0.450 |
| <30 days | 7 | 6 | 4 |  |
| ≥ 30 days | 6 | 6 | 9 |  |
| Smoking |  |  |  | 0.648 |
| Never smoker | 4 | 3 | 2 |  |
| Former/current smoker | 9 | 9 | 11 |  |
| *Tumor burden* |  |  |  |  |
| Median RECIST score | 76 | 91 | 94 | 0.798 |
| Median bMAF (%) | 3.3 | 9.0 | 3.4 | 0.828 |

### Differences in TMB among sites of tissue biopsy

To assess whether there was any systematic bias towards TMB levels according to the site of tissue biopsy, TMB levels were compared across biopsy sites (Figure 2). Median tTMB and bTMB values in patients whose tissue was sampled from lymph nodes were higher than those from either primary or metastatic lesions, and there was a significant difference of distribution in tTMB (Kruskal-Wallis test, p = 0.002) but not in bTMB (p = 0.058).

**Figure 2.**
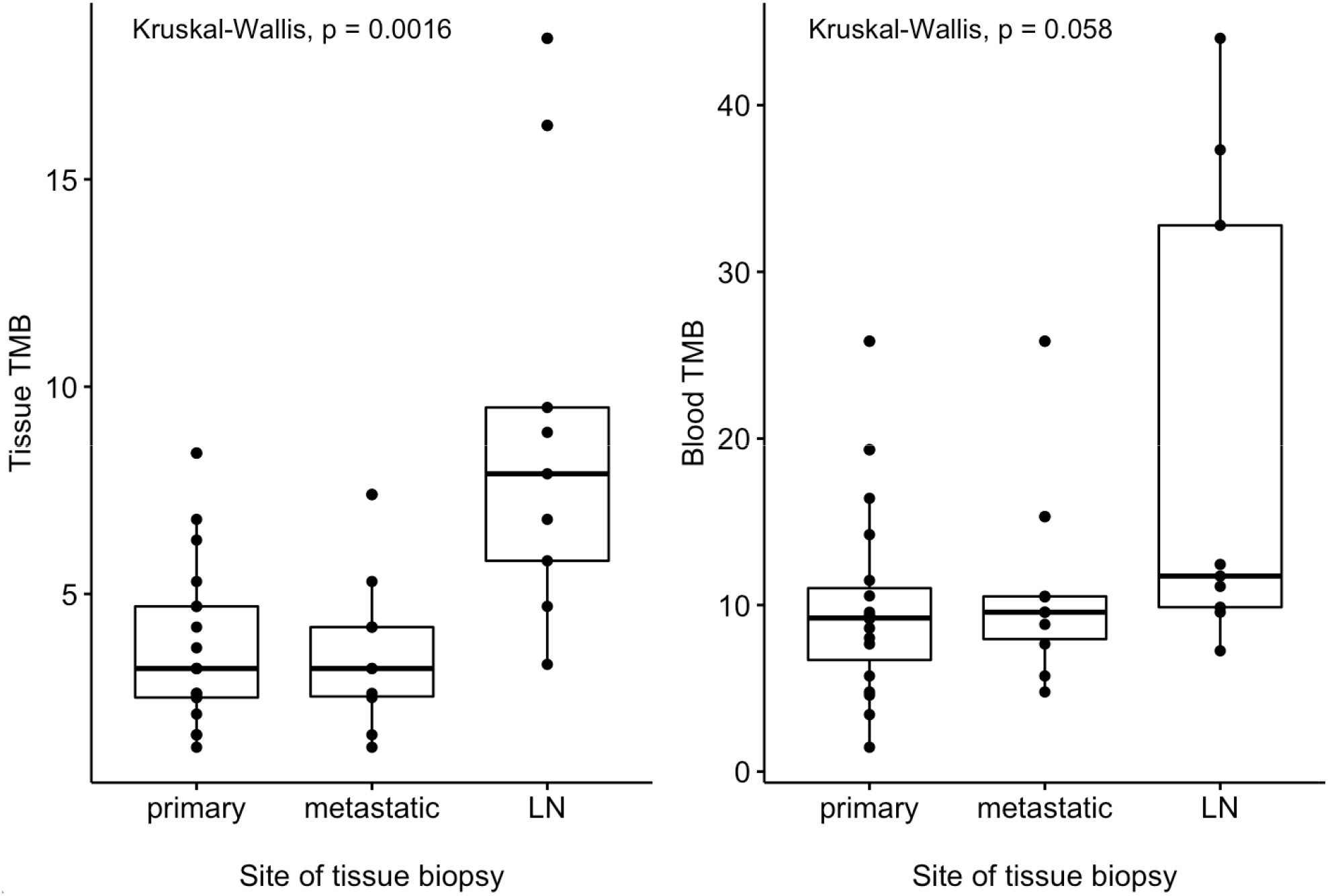
Boxplot distribution of tissue or blood tumor mutational burden by site of tissue biopsy Tissue tumor mutational burden of patients whose tissue was sampled from lymph nodes was higher than those whose tissue was sampled from primary lesions or metastatic lesions (excluding lymph nodes). TMB, tumor mutational burden; LN, lymph nodes.

## Discussion

Our current study attempted to delineate the concordance between bTMB and tTMB as well as identify potential patient characteristics and clinical variables that might influence concordance. For the first time, we used the two commonly used commercial platforms of measuring TMB to identify the ratio of bTMB to tTMB in real-world settings and explore potential factors that could contribute to any discordance between bTMB and tTMB.

In line with a previous large-scale study, bTMB showed a moderate level of correlation with tTMB and bTMB levels were higher than tTMB levels.^6^ Of note, while only two patients had tTMB ≥ 10 mut/Mb, 16 patients had bTMB ≥ 10 mut/Mb, and bTMB:tTMB ratio at 10 mut/Mb tTMB was 2.1:1. Consistent reports of higher TMB in blood than in tissue suggests that using bTMB of ≥ 10 mut/Mb as a threshold could result in unnecessary treatment, especially since only a minority of immunogenic patients respond to immunotherapy. In addition, some patients receiving immunotherapy experience resistance, serious immune-related adverse events, or even accelerated disease progression, also known as hyperprogression.^14^

Exactly how much bTMB is higher than tTMB and the ramifications of this difference to patient care remains to be further verified. The bTMB:tTMB ratio of 2.1:1 at tTMB of 10 mut/MB reported in our study is higher than 1.6:1 reported in the MYSTIC trial, but the median bTMB reported in our study is lower (9.6 vs. 13.4 mut/Mb).^15^ Among many plausible causes, this difference could be attributed to the differences in assays; while blood TMB was measured using the same platform, tissue TMB was measured with FoundationOne in the MYSTIC trial. As the technology of TMB calculation is relatively new and rapidly changing, significant variability exists between TMB levels calculated by different assays, and harmonization of TMB levels across these assays will be important in defining the concordance between bTMB and tTMB.^9^

We also found that stratification of patients by cancer type and site of tissue biopsy could alter the degree of correlation significantly. Higher correlation between bTMB and tTMB was observed in tumors sampled from metastatic lesions (excluding lymph nodes) or in non-lung cancers. This observation in metastatic lesions is plausible, since metastatic tissue tend to have a higher TMB than primary tissue, possibly due to the increased clonality of cells with high mutational burden that metastasize, also known as “bottlenecking”; since bTMB is usually higher than overall tTMB, it will correlate better with higher tTMB values.^13^ While it is unclear why higher correlation was observed for non-lung cancers, it could partly be due to the fact that many of our patients with non-lung cancers had their tissue biopsy taken from metastatic lesions.

Classifying patient TMB levels as concordant or discordant based on bTMB:tTMB ratio revealed some patterns, but no significant associations with any of the patient characteristics or tumor burden. tTMB reflects the mutational burden of a single site in a tumor, whereas bTMB is derived from all cancer cells that release DNA into the blood and could therefore be more representative of systemic tumor burden.^10^ MAF is associated with the amount of circulating tumor DNA (ctDNA) in the blood and with lower overall survival, and was thus used as a molecular marker of tumor burden in our study.^10, 11^ However, neither RECIST score nor MAF was different between patients with concordant and discordant bTMB:tTMB ratios. The current findings suggest that bTMB levels may be independent of tumor burden, and that there may be other causes of discordance between bTMB and tTMB that demand further exploration. One interesting finding from our analysis was that both tTMB and bTMB was higher in patients whose tissue was sampled from lymph nodes, although the statistical association in bTMB was marginally not significant. In contrast, previous studies have reported association between lower TMB and lymph node metastasis in breast and colon cancer, citing immune evasion as a possible mechanism.^16, 17^ In addition to considering molecular explanations, it may be important to incorporate the clinical picture, as lymph nodes are more likely to be biopsied if regional metastasis is suspected, or are simply large enough to provide sufficient tissue. While further studies are required, these results highlight the often-overlooked implication of the site of tumor biopsy.

To our knowledge, this is the first study to use TMB values reported on two different commercially available assays to explore concordance between tissue and blood TMB. Our study has several limitations. First, the relatively small number of patients may have led to coincidental findings, or on the contrary, inability to discover significant relationships. Second, our study population had a heterogeneous distribution of cancer types with a bias towards lung cancer, but this in turn allowed us to reveal differing patterns of bTMB:tTMB concordance between lung cancer and other types of cancer.

## Conclusion

bTMB was positively correlated with tTMB, and median bTMB was higher than median tTMB. Cancer type and site of tissue biopsy may influence concordance between tTMB and bTMB. Future studies with more patients may help define the optimal bTMB threshold for receiving immunotherapy, which may be different from the tTMB threshold.

## Data Availability

None

## Acknowledgements

We would like to thank Leslie Kiedrowski for her opinion on interpreting tumor mutational burden levels.

## Notes

### Funding Statement

No funding was received

### Author Declarations

The study was approved by the Institutional Review Board (IRB) of Northwestern University Feinberg School of Medicine. Since retrospective review of molecular analyses was performed, informed consent was waived.

## References

1. Goodman AM, Kato S, Bazhenova L, et al: Tumor mutational burden as an independent predictor of response to immunotherapy in diverse cancers. Mol Cancer Ther 16:2598–2608, 2017

2. Subbiah V, Solit DB, Chan TA, et al: The FDA approval of pembrolizumab for adult and pediatric patients with tumor mutational burden (TMB) ≥10: a decision centered on empowering patients and their physicians. Ann Oncol 31:1115–1118, 2020

3. Fenizia F, Pasquale R, Roma C, et al: Measuring tumor mutation burden in non-small cell lung cancer: Tissue versus liquid biopsy. Transl Lung Cancer Res 7:668–677, 2018

4. Gandara DR, Paul SM, Kowanetz M, et al: Blood-based tumor mutational burden as a predictor of clinical benefit in non-small-cell lung cancer patients treated with atezolizumab [Internet]. Nat Med 24, 2018 Available from: http://dx.doi.org/10.1038/s41591-018-0134-3

5. Aggarwal C, Thompson JC, Chien AL, et al: Baseline Plasma Tumor Mutation Burden Predicts Response to Pembrolizumab-based Therapy in Patients with Metastatic Non–Small Cell Lung Cancer. Clin Cancer Res 26:2354–2361, 2020

6. Rizvi NA, Cho BC, Reinmuth N, et al: Durvalumab with or Without Tremelimumab vs Standard Chemotherapy in First-line Treatment of Metastatic Non-Small Cell Lung Cancer: The MYSTIC Phase 3 Randomized Clinical Trial [Internet]. JAMA Oncol 6:661–674, 2020 Available from: https://jamanetwork.com/journals/jamaoncology/fullarticle/2763864

7. Davis AA, Chae YK, Agte S, et al: Comparison of tumor mutational burden (TMB) across tumor tissue and circulating tumor DNA (ctDNA). [Internet]. J Clin Oncol 35:e23028–e23028, 2017 Available from: https://doi.org/10.1200/JCO.2017.35.15_suppl.e23028

8. Chae YK, Davis AA, Agte S, et al: Clinical Implications of Circulating Tumor DNA Tumor Mutational Burden (ctDNA TMB) in Non-Small Cell Lung Cancer [Internet]. Oncologist 24:820–828, 2019 Available from: https://pubmed.ncbi.nlm.nih.gov/30867242

9. Merino DM, McShane LM, Fabrizio D, et al: Establishing guidelines to harmonize tumor mutational burden (TMB): in silico assessment of variation in TMB quantification across diagnostic platforms: phase I of the Friends of Cancer Research TMB Harmonization Project [Internet]. J Immunother Cancer 8:e000147, 2020 Available from: https://jitc.bmj.com/lookup/doi/10.1136/jitc-2019-000147

10. Chae YK, Davis AA, Agte S, et al: Clinical Implications of Circulating Tumor DNA Tumor Mutational Burden (ctDNA TMB) in Non□Small Cell Lung Cancer [Internet]. Oncologist 24:820–828, 2019 Available from: https://onlinelibrary.wiley.com/doi/abs/10.1634/theoncologist.2018-0433

11. Wang Z, Duan J, Wang G, et al: Allele Frequency–Adjusted Blood-Based Tumor Mutational Burden as a Predictor of Overall Survival for Patients With NSCLC Treated With PD-(L)1 Inhibitors [Internet]. J Thorac Oncol 15:556–567, 2020 Available from: https://doi.org/10.1016/j.jtho.2019.12.001

12. Yang N, Li Y, Liu Z, et al: The characteristics of ctDNA reveal the high complexity in matching the corresponding tumor tissues. BMC Cancer 18:1–12, 2018

13. Schnidrig D, Turajlic S, Litchfield K: Tumour mutational burden: primary versus metastatic tissue creates systematic bias. Immuno-Oncology Technol 4:8–14, 2019

14. Fountzilas E, Kurzrock R, Hiep Vo H, et al: Wedding of Molecular Alterations and Immune Checkpoint Blockade: Genomics as a Matchmaker. JNCI J Natl Cancer Inst, 2021

15. Si H, Kuziora M, Quinn KJ, et al: A blood-based assay for assessment of tumor mutational burden in first-line metastatic NSCLC treatment: Results from the MYSTIC study A C. Clin Cancer Res 27:1631–1640, 2021

16. Wang Z, Liu W, Chen C, et al: Low mutation and neoantigen burden and fewer effector tumor infiltrating lymphocytes correlate with breast cancer metastasization to lymph nodes. Sci Rep 9:1–10, 2019

17. Huang X, Cai W, Liu L, et al: Low mutation burden and differential tumor-infiltrating immune cells correlate with lymph node metastasis in colorectal cancer. Int J Clin Exp Pathol 13:2259–2269, 2020

